# Usability, Safety and Tolerability of CUE1 Vibrotactile Device as Promising Therapeutic Approach in Orthostatic Tremor

**DOI:** 10.1101/2025.07.23.25332038

**Authors:** Viktoria Azoidou, Alexandra Zirra, Thomas Boyle, David Gallagher, Alastair John Noyce, Cristina Simonet

**Author notes:** Corresponding author: Viktoria Azoidou, Centre for Preventive Neurology, Wolfson Institute of Preventive Medicine, London, United Kingdom. &.

## Abstract

**Introduction:** Primary Orthostatic Tremor (POT) is a rare hyperkinetic movement disorder characterized by unsteadiness while standing, often exacerbated by anxiety and fatigue. It significantly impairs quality of life, and current treatment options are limited, with invasive procedures carrying notable risks.

**Methods:** This unblinded interventional study assessed the usability, safety, and tolerability of the CUE1, a non-invasive vibrotactile stimulation device worn on the sternum. Secondary exploratory outcomes included objective measures of balance and mobility: maximal stance time, tandem walk, heel raise hold, tandem stance, and Timed Up and Go (TUG) with and without dual tasking (DT). Assessments were performed at baseline, immediately after a 20-minute acclimatization session, and after 9 weeks of daily use (8 hours/day). Patient-reported outcomes on fatigue, anxiety, and perceived change were also collected.

**Results:** Ten participants with POT (70% female, aged 58-88 years) completed the study. Compliance and tolerability were excellent (100%), with only mild, transient skin irritation reported in two cases. Immediate post-intervention improvements were observed in TUG [-1.22seconds (95% CI: -2.03, -0.26), p=0.020], tandem stance [+1.41seconds (0.00, 6.13), p=0.025], tandem walk [+0.50 steps (0.00, 2.50), p=0.042], and heel raise hold [+1.62seconds (1.00, 3.32), p=0.020]. After 9 weeks, improvements were observed in TUG [-2.13seconds (−4.00, -0.26), p=0.028], TUG-DT [-7.51seconds (−14.88, -0.14), p=0.047], tandem stance [+9.06seconds (1.04, 17.08), p=0.028], fatigue [-7.00 (−13.63, -0.37), p=0.035], and patient-reported impression of change [+1.10 (0.14, 2.06), p=0.027].

**Conclusion:** CUE1 vibrotactile stimulation is safe, well-tolerated, and shows promise in improving balance, mobility, and fatigue in POT. Larger, controlled trials are warranted.

**HIGHLIGHTS:**

- This pilot interventional study evaluated 9 weeks of daily vibrotactile stimulation using the non-invasive CUE1 device in patients with Primary Orthostatic Tremor (POT).
- All ten participants completed the study with 100% compliance and no serious adverse events; only mild, temporary skin irritation was reported.
- Immediate improvements were seen in balance and mobility measures after a single 20-minute session with the device.
- Continued use over 9 weeks led to significant gains in TUG (with and without dual tasking), tandem stance, and reduced fatigue.
- The CUE1 was safe, well-tolerated, and shows potential as a non-invasive therapeutic option for POT; however, large controlled trials are needed.

## INTRODUCTION

Primary orthostatic tremor (POT) is a rare hyperkinetic disorder marked by high-frequency tremors primarily in the legs, trunk, and occasionally the arms, predominantly when standing still[1,2]. The exact prevalence of the condition remains undetermined; however, numerous cases have been documented globally, with a higher incidence observed in females (sex ratio: 2:1). The average age of onset is approximately 55 to 60 years, with a reported range spanning from 13 to 85 years[1,2].

POT leads to significant postural unsteadiness, and often accompanied by fatigue, anxiety, and psychological distress related to poor balance[3]. Gait abnormalities, such as reduced speed and impaired tandem walking, may also occur[4]. While early neurophysiological studies identified a tremor frequency of 13-18 Hz (e.g., fast POT), more recent research suggests that tremor frequencies may be below 10 Hz (e.g., slow POT) in some cases[1]. POT severely impacts daily activities such as cooking, showering, and standing in line, as the tremors primarily occur when standing still. Over time, the condition often progresses, leading to increasing disability, significantly affecting quality of life (QoL)[3,4].

The pathophysiology of POT suggests a central origin primarily in the brainstem and cerebellum, supported by observations like cerebellar atrophy, abnormal posturography, and bilateral tremor coherence[1,2]. Additionally, dopaminergic dysfunction and involvement of multiple brain regions, including the basal ganglia and motor cortex, may contribute to POT generation. Currently, there is no cure for POT. Clonazepam is the first line treatment for POT with at least mild benefit seen in about half of the cases[1]. Second line treatment include other benzodiazepines (e.g. diazepam, lorazepam), beta-blockers, and antiepileptic drugs. Side effects or lack of response are the main reason of discontinuation of this treatment. Surgical interventions such as deep brain stimulation (DBS) targeting the bilateral thalamic ventral intermediate nuclei and spinal cord stimulation (SCS) have shown promise in small cohorts [1,5], but their clinical application is limited due to potential severe side effects including balance problems and persistent paraesthesia[5]. Despite available treatments, the effectiveness of current approaches for POT remains limited, underscoring the need for novel, non-invasive therapies. While targeting tremor and balance issues may offer benefit, the impact on related symptoms such as fatigue and anxiety is even less unclear. Exploring these symptoms is also essential to fully understand the broader effects of POT. Cueing is a non-invasive technique using external prompts (e.g., visual, auditory and proprioceptive) to aid motor function. It has shown to be safe and well tolerated and possibly effective in improving motor symptoms and non-motor symptoms in people with Parkinson’s disease (PwP)[6,7]. Focused vibrotactile stimulation, which delivers localized skin vibrations through non-invasive devices like vibrotactile socks[6] or CUE1 device[7], has also been shown to enhance balance, and gait in PwP, with potential applications for tremor control in PwP[7]. Similarly, it may be effective in individuals with essential tremor [8].Potential effects on POT have not been explored. The primary aim of this unblinded 9-week interventional study was to assess the usability, safety and tolerability of CUE1 in POT. The second aim was to explore the effects of CUE1 on tremor-related balance difficulties in people with POT, with a focus also on associated symptoms such as anxiety and fatigue.

## METHODS

### Trial Design

This 9-week unblinded interventional study was conducted at Queen Mary University of London (QMUL), United Kingdom (UK) (reference number: 23/PR/1526; clinical.trials.gov ID: NCT06174948). The study adhered to the Declaration of Helsinki. All participants provided written informed consent before data collection. Participants were identified from the Neurology Department at Barts Health NHS Trust, the Department of Care of the Elderly at Homerton Healthcare NHS Foundation Trust, and the Orthostatic Tremor UK Support Group. Recruitment and all assessments were conducted at the Centre for Preventive Neurology, QMUL.

### Participants

Inclusion criteria consisted of a clinical examination and diagnosis of POT[8] confirmed by electromyography (EMG), participants aged over 18 years, and the ability to provide written informed consent. Exclusion criteria included alternative diagnoses affecting movement or balance (e.g., Parkinson’s disease, atypical parkinsonism, cerebellar ataxia), other tremor types (e.g., essential tremor, titubation, functional tremor, orthostatic myoclonus), osteoarticular disorders, sensory disturbances, orthostatic hypotension or other cardiovascular causes of postural intolerance, cognitive impairments (e.g., Alzheimer’s), or started other treatments for POT less than 3 months ago. Additionally, participants with implanted metallic/electronic devices, hypersensitivity to vibrotactile stimulation, or skin conditions near the device application site were excluded.

### Measures

Baseline demographic data including age, sex, ethnicity, disease duration from diagnosis confirmation, medication, and cognitive status (Montreal Cognitive Assessment tool; MoCA) were collected.

The primary outcome was home usability of CUE1 over 9-weeks, assessed via recruitment, adherence, dropout rates, and safety/tolerability. Site-level usability included engagement metrics, and protocol compliance was tracked through timely intervention delivery, consistent assessments, and complete data. Investigators documented participant flow (screening, eligibility, approach, consent, enrolment). Participant-reported usability covered adherence, retention, and completeness of baseline and follow-up data. Compliance was measured using (1) a 5-point Likert scale for frequency of use (0=not at all; 4=daily ≥8 hours) and (2) a 0-10 scale for perceived compliance. Safety and tolerability were monitored throughout, with all adverse events and malfunctions recorded. At study end, participants rated satisfaction on a 5-point Likert scale.

Participants underwent two in-person assessments: one at baseline and one at the 9-week follow-up to measure the exploratory effectiveness outcomes. At the baseline visit, each participant was first assessed without the CUE1 device, followed by a repeat of the same assessments after a 20-minute acclimatization period wearing the CUE1. As per prior protocols and company guidance, the device must be worn for at least 20 minutes before any potential benefits can be observed[6]. At the 9-week follow p assessments, participants were assessed without the CUE1 on them. The main comparison was made between the initial baseline scores (device off) and those recorded at the 9-week follow-up (also device off). This 9-week comparison was designed specifically to minimize short-term learning effects, as this assessment was performed only once at that session and spaced far apart in time from baseline assessment.

Exploratory effectiveness outcomes included balance and gait assessments: maximal stance time with feet together and apart, maximal stance time during tandem stance, tandem walk, heel raise hold and Timed Up and Go (TUG) with and without a dual task (DT), which involved counting backwards by sevens from 100. All balance and gait measures are reported in seconds, except for the tandem walk, which is recorded as the maximum number of consecutive steps taken. Patient-reported outcomes included the Fatigue Severity Scale (FSS), Hospital Anxiety and Depression Scale (HADS), and Patient Global Impression of Change (PGIC).

### Intervention

Participants used the CUE1 daily at home while continuing their usual activities. The CUE1 device (Charco Neurotech Ltd, UK) is a CE-marked, non-invasive neuromodulation aid for managing motor symptoms in Parkinson’s. It is worn on the sternum via an adhesive patch and delivers high-frequency vibration (80-250 Hz) and low-frequency cueing (pulse and rest length between 100-2000 ms). Participants used a fixed stimulation protocol (80% intensity, 800 ms pulse/rest of the above range) without access to the CUE App (e.g., gives the opportunity to modify the stimulation parameters) to ensure standardization. The 9-week intervention duration was based on prior studies in PwP[6] (Supplementary Material A for device details). The baseline intervention involved wearing the CUE1 for 20 minutes prior to testing to assess its initial effects. Following this assessment session, participants wore the device for 8 continuous hours daily over a 9-week period, starting each morning. Participants were asked to continue taking their regular medication throughout the duration of the trial.

### Statistical Analysis

Statistical analyses were performed using IBM SPSS Statistics (version 29; IBM Corp., Armonk, NY, USA). Descriptive statistics summarised baseline and outcome data. Normally distributed continuous variables are reported as mean (SD), and non-normal data as median (IQR). Categorical variables are presented as frequency (n) and percentage (%). Normality was assessed via Shapiro-Wilk test, supported by histogram and Q-Q plot inspection. Ninety-five percent confidence intervals (95% CI) are provided where relevant. Secondary effectiveness outcomes were exploratory, with significance set at p<0.05. Within-group comparisons used paired t-tests or Wilcoxon signed-rank tests, as appropriate. Non-parametric methods were applied when distributions differed between timepoints.

## RESULTS

The trial was completed as planned, with no delays or protocol changes. Between March 29 and December 11, 2024, 92 individuals were screened: 85 from the Orthostatic Tremor UK Support Group, 3 from Homerton University Hospital, and 4 from Barts Health NHS Trust. Most were excluded due to distance from the site (n=76), lack of confirmed diagnosis (n=5), or co-existing conditions affecting balance (n=1) (Supplementary Material B). Ten participants with POT were enrolled (30% male, 70% female) (Table 1). Four (40%) were on stable medication, most commonly gabapentin, for at least three months, with no changes during the study.

**Table 1.** Participants’ demographic and clinical characteristics.

| Demographic Characteristics | Participants (n = 10) |
| --- | --- |
| Age (years), mean ± SD | 69.90 ± 8.56 |
| Gender |  |
| Male, n (%) | 3 (30%) |
| Female, n (%) | 7 (70%) |
| Ethnicity |  |
| White or White British (n, %) | 9 (90%) |
| Asian or Asian British (n, %) | 1 (10%) |
| Diagnosis, n (%) |  |
| Primary Orthostatic Tremor |  |
| Slow (<10 Hz) | 2 (20%) |
| Fast (13-18 Hz) | 8 (80%) |
| Symptom duration (years), mean ± SD | 10.30 ± 9.68 |
| Medication for Primary Orthostatic Tremor |  |
| Clonazepam | 1 (10%) |
| Gabapentin | 2 (20%) |
| Propranolol | 1 (10%) |
| None | 6 (60%) |
| MoCA, mean ± SD | 27.00 ± 1.89 |

Data are n (%) or mean (SD). N= number. % = Percentage. SD = Standard deviation. MoCA, Montreal Cognitive Assessment tool.

Recruitment was low (11%), likely due to the rarity of POT, but there were no dropouts. All 10 participants completed the 9-week intervention, wearing the CUE1 device for approximately 500 hours each. No adverse reactions or device malfunctions were reported. Two participants experienced mild skin irritation, which was resolved by adjusting the adhesive patch location. No difficulties were reported with patch application or removal. Tolerability was excellent, with 100% compliance and high participant-reported usability (Table 2a). Overall, the CUE1 device was safe, well-tolerated, and consistently used throughout the study period. Participants’ feedback was uniformly positive, with all expressing interest in continued use of CUE1 post-trial and willingness to recommend the device to others (Supplementary Material C).

**Table 2.**
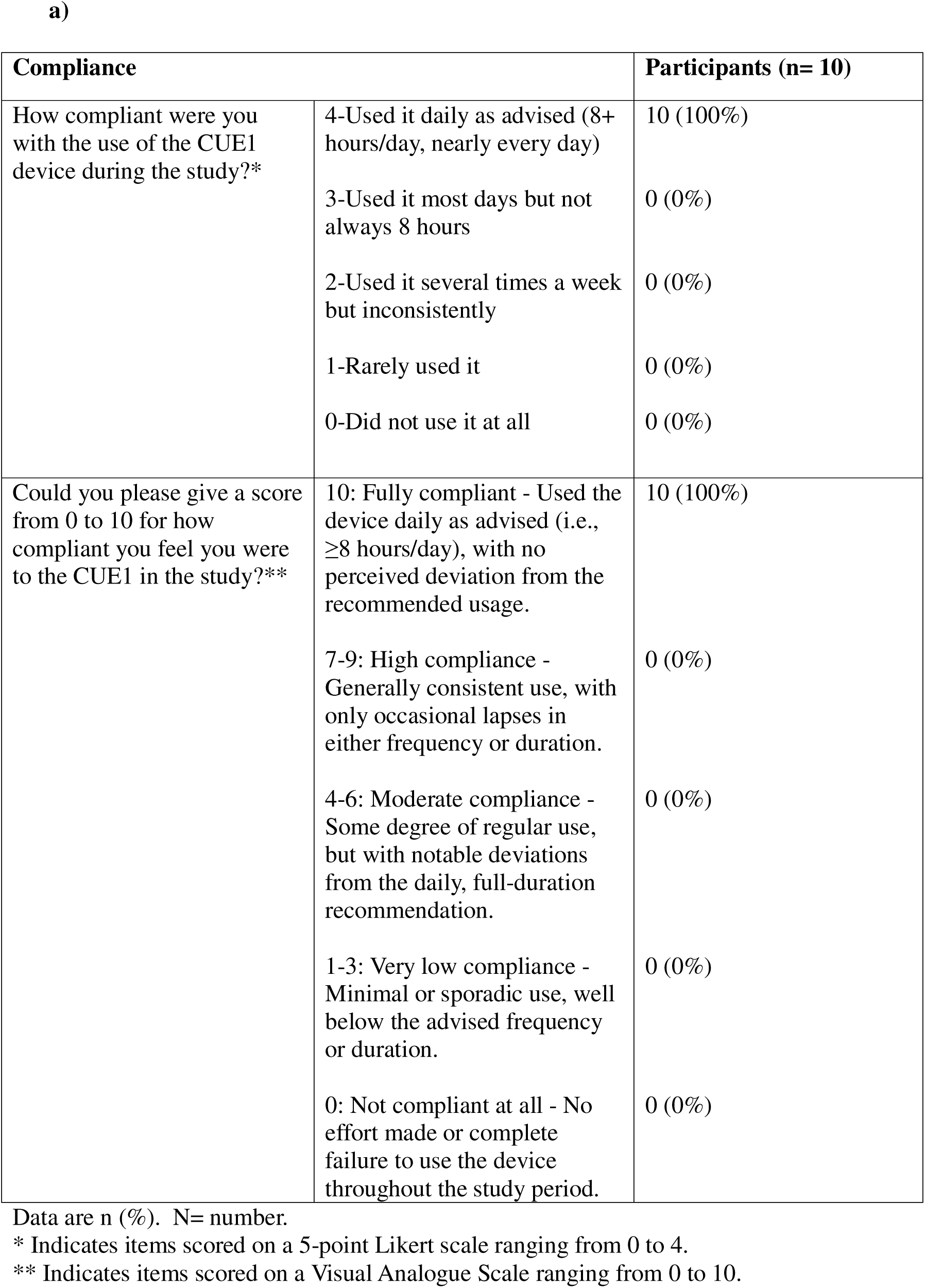
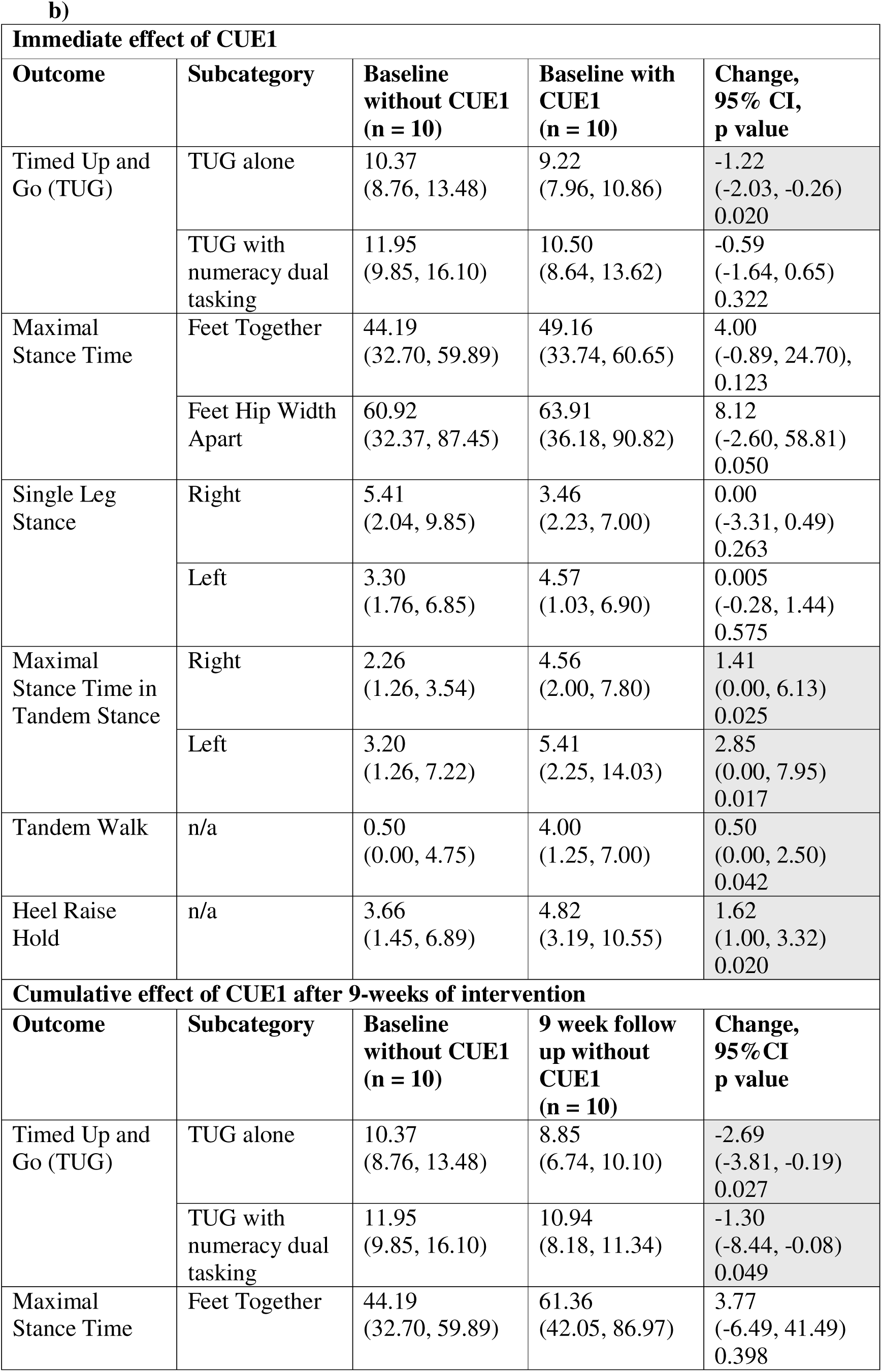
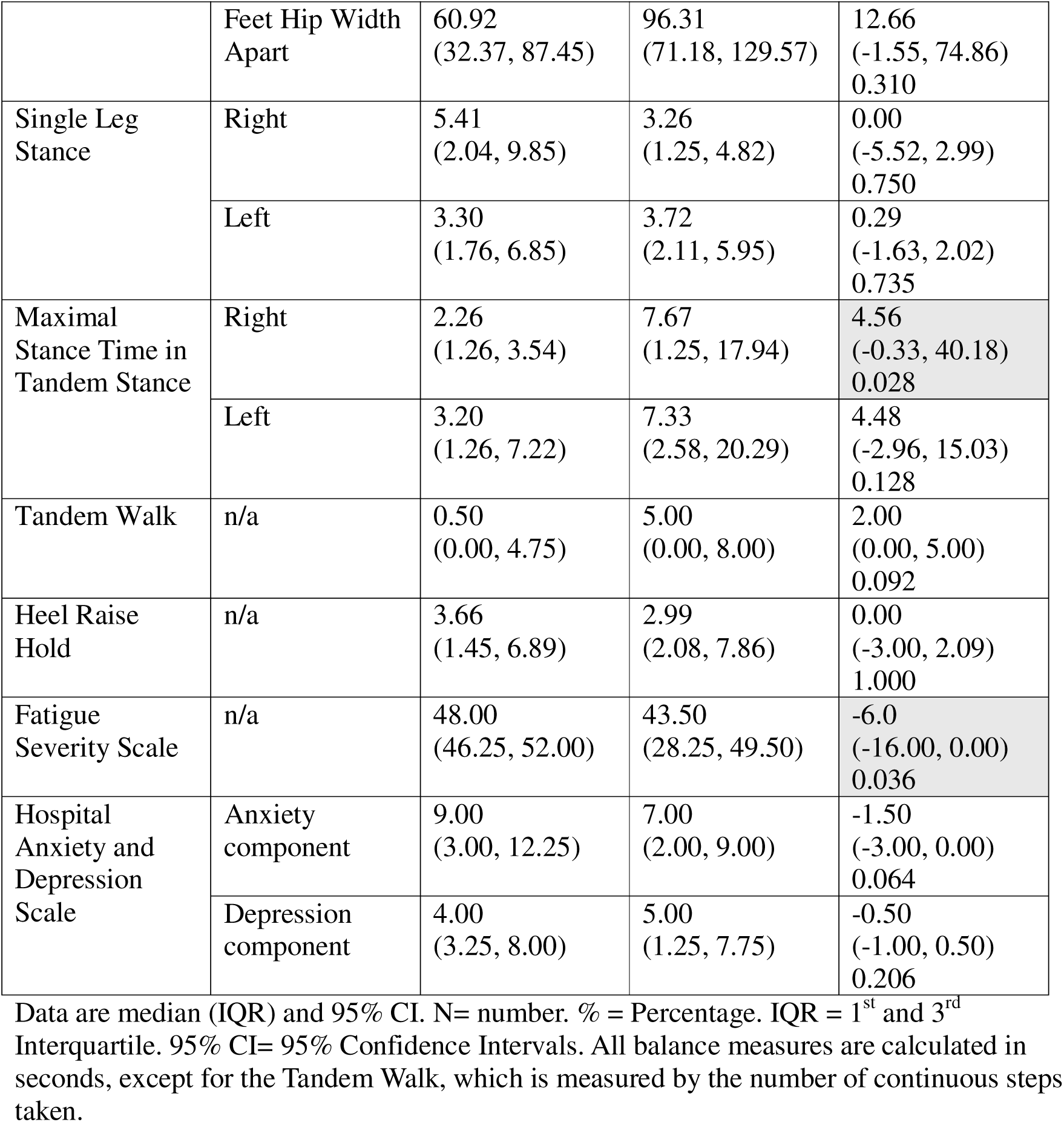
Table 2a shows patient-reported compliance with the CUE1 intervention, while Table 2b presents the immediate and cumulative effects of CUE1 over the 9-week intervention period.

Participants demonstrated immediate improvements in several functional measures following the intervention (Table 2b). Gains were observed in TUG performance, tandem stance duration, tandem walk steps, and heel raise hold. At the 9-week follow-up, further improvements were noted in TUG, TUG DT, and tandem stance. Fatigue scores also improved significantly. Participants indicating improvement on the Global Impression of Change [4 (2-5)] and Degree of Change [3.90 (1.45)]. However, it is important to note that the effectiveness outcomes were exploratory in nature, and the study was not powered to detect definitive treatment effects.

## DISCUSSION

This study is the first to explore the potential effects of non-invasive vibrotactile stimulation with cueing on postural control in patients with POT. Currently, there are no established long-term non-invasive treatment options for POT. Previous research has focused on interventions such as theta burst repetitive transcranial magnetic stimulation, which demonstrated short-term improvements in postural stability and tremor intensity, though it did not alter tremor frequency [10]. Similarly, proprioceptive soleus muscle tendon vibration has been associated with temporary improvements in postural control and tremor intensity, without affecting tremor frequency [11].

In our study, we observed trends suggestive of short-term changes in balance and gait following use of the CUE1 device, with some participants showing effects persisting up to 9-weeks. Although these changes did not reach clinical significance, they raise the possibility that vibrotactile cueing could influence postural control in POT. Potential mechanisms may include modulation of tremor amplitude or alterations in proprioceptive processing[6,7,10,11]. However, given the uncontrolled nature of the study and the small sample size, these preliminary findings should be interpreted with caution. Further controlled studies are needed to clarify whether the observed changes are attributable to the intervention or to other factors such as placebo effects, natural symptom fluctuations, or increased patient awareness.

Knowledge gaps remain regarding tremor discharge mechanisms and postural alterations in POT[1]. A ponto-cerebello-thalamo-cortical network is suggested as a pathophysiological correlate [4,14]. The role of proprioceptive feedback and effect of vibrotactile cueing on postural control in POT is unclear, requiring further investigation on balance and gait outcomes. Previous studies show disproportionate increases in postural instability relative to tremor severity[10,11]. It is possible that different stimulation frequencies could influence tremor severity differently in POT and this should be investigated. For instance, 80 Hz vibrotactile devices showed no significant changes in tremor amplitude or stability in PwP[6,11], while a 250 Hz device showed promise in Essential Tremor[8].

Vibrotactile stimulation has emerged as a promising non-invasive approach to improve balance and gait, though current evidence in PwP is limited to underpowered studies[6,7]. Given the lack of research on non-invasive therapies in patients with POT, further investigation on vibrotactile stimulation is warranted. Exploring the impact of different stimulation frequencies may optimize therapeutic outcomes by reducing postural instability, alleviating fear of falling, and enhancing QoL. Accordingly, we evaluated the effects of vibrotactile stimulation on balance and gait through ecologically valid, functional tasks such as walking with DT, which are often impaired in POT[4], rather than relying solely on tremor metrics[1,2]. We observed patterns suggestive of both short-term and cumulative improvements in balance and gait, with and without DT, that are broadly consistent with previous findings on the use of CUE1 and other vibrotactile devices in PwP [7,13]. Although POT is primarily characterized by postural instability, our preliminary data indicate that functional mobility, as assessed by the TUG test may also be responsive to intervention. In this small, uncontrolled cohort, median TUG times improved immediately after 20 minutes of CUE1 use and again after 9 weeks of daily wear. However, given the lack of a control group and potential for learning effects, these trends should be interpreted with caution. Nonetheless, they suggest that CUE1 may have the potential to support functional outcomes in individuals with POT, warranting further investigation in larger, controlled studies.

Patients with POT face significant challenges in daily activities and social interactions, often avoiding situations that require standing still, leading to physical and mental exhaustion[3]. Fatigue is common in POT, yet no studies have specifically assessed it following interventions. Our study is the first to evaluate fatigue in response to non-invasive treatment. Fatigue improvements may result from enhanced postural stability which may have freed cognitive resources, reducing fatigue. Additionally, enhanced stability may have contributed to increased engagement in daily activities, potentially further reducing fatigue. However, the impact of this increased activity depends on the nature of the tasks; if the activities are physically demanding, they could, in fact, exacerbate fatigue.

This study highlights the potential of vibrotactile stimulation in patients with POT; however, several limitations must be acknowledged. The uncontrolled design, lack of blinding, and small sample size limit the strength and generalizability of the findings. Tremor characteristics were not directly measured as outcomes, making it unclear whether observed changes in balance and gait were linked to tremor modulation or other mechanisms, such as altered proprioception. The use of a fixed stimulation frequency and placement may have constrained the impact of the intervention. Exploring alternative parameters and anatomical targets such as the lower back or hips may be beneficial in future studies. Additionally, postural instability as a surrogate for tremor requires further validation. More comprehensive assessments, including both quantitative and qualitative measures of QoL, are particularly important in rare conditions like POT[1,3,9]. A multidisciplinary approach may help optimise treatment strategies and patient outcomes.

## CONCLUSIONS

This study provides preliminary evidence that cueing with focused vibrotactile stimulation using the CUE1 is a feasible, safe, and well-tolerated treatment for patients with POT. It shows promise in improving balance and reducing fatigue. Double-blind control trials are needed to confirm these findings.

## Data Availability

All data produced in the present study are available upon reasonable request to the authors.

## ACKNOWLEDGEMENTS

We would like to thank all participants for taking part in this study and Orthostatic Tremor UK Support Group for their support. We appreciate the efforts of all the authors for this article. This study is part of the project funded by Knowledge Transfer Partnership (KTP) United Kingdom 2021 to 2022, round 4, UKRI KTP (Innovate UK) and we would like to thank the organisation for the funding.

## DATA STATEMENT

The datasets generated during and/or analyzed during the current study are available upon reasonable request from the corresponding author.

## DECLARATION OF INTERST STATEMENT

### Financial disclosures for the previous 12 months

Prof Noyce reports grants from Parkinson’s UK, Barts Charity, Cure Parkinson’s, National Institute for Health and Care Research, Innovate UK, Virginia Keiley benefaction, Solvemed, the Medical College of Saint Bartholomew’s Hospital Trust, Alchemab, Aligning Science Across Parkinson’s Global Parkinson’s Genetics Program (ASAP-GP2) and the Michael J Fox Foundation. Prof Noyce reports consultancy and personal fees from AstraZeneca, AbbVie, Profile, Bial, Charco Neurotech, Alchemab, Sosei Heptares, Umedeor and Britannia, outside the submitted work. Prof Noyce has share options in Umedeor. Prof Noyce is an Associate Editor for the Journal of Parkinson’s Disease.

Dr Cristina Simonet reports grants from Innovate UK and the Michael J Fox Foundation. She also works as a co-investigator with Roche, outside the submitted work.

## ETHICAL COMPLIANCE STATEMENT

The datasets generated during and/or analyzed during the current study are available upon reasonable request from the corresponding authors.

This study was approved in the UK by the Dulwich Research Ethics Committee (reference number: 23/PR/1526; clinical.trials.gov ID: NCT06174948). The study adhered to the Declaration of Helsinki. All patients provided written informed consent before data collection.

We confirm that we have read the position of the Journal on issues involved in ethical publication and affirm that this work is consistent with those guidelines.

**Supplemental material A.**
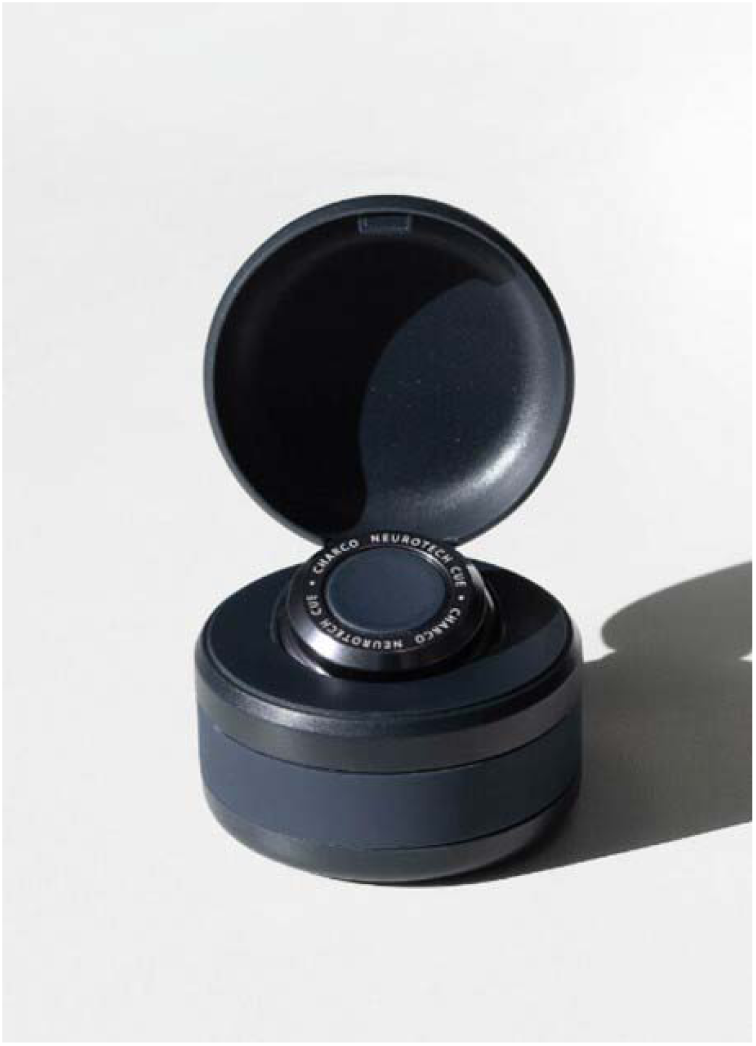
The CUE1 device. The CUE1 device is designed to provide non-invasive support for both motor and non-motor symptoms of Parkinson’s. It is a compact device, with dimensions of 40 mm in diameter, 11 mm in height, and a weight of 17 g. The device attaches to the sternum using dermatologically tested adhesive patches that are waterproof, with each patch capable of remaining in place for up to 14 days without replacement. While the CUE1 itself is water-resistant, it is not fully waterproof and should be removed prior to exposure to water, such as during showering. The CUE1 utilizes a silent motor to deliver vibrotactile stimulation, with a stimulation pattern developed through user testing in people with Parkinson’s and Primary Orthostatic Tremor. Although initial clinical trials have been conducted in people with Parkinson’s, clinical trials are needed to assess the mechanisms, feasibility, safety, tolerability, and efficacy of CUE1 in people with Primary Orthostatic Tremor.

**Supplemental material B.**
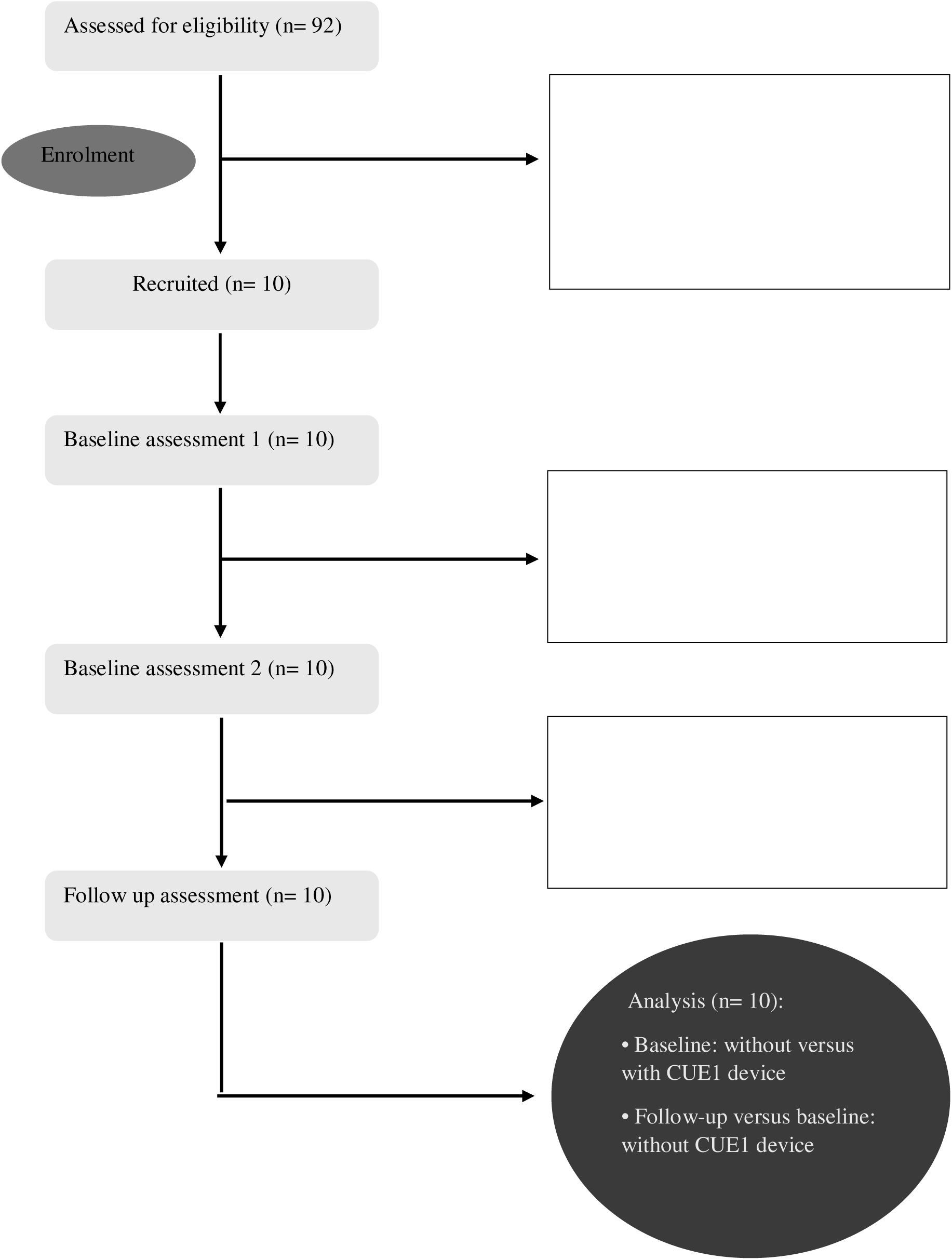
Flow of participants through this study.

**Supplemental material C.** Participants’ feedback on receiving CUE1 intervention.

| **Participants' Satisfaction with CUE1** | **Participants**<br>**(n = 10)** |
| --- | --- |
| How helpful did you find the CUE1 device in alleviating your symptoms?<sup>a</sup> | 1.55 (0.93) |
| How satisfied were you with using the CUE1 device?<sup>a</sup> | 3.00<br>(1.00, 3.00) |
| If given the option, how likely are you to continue using the CUE1 device?<sup>a</sup> | 3.00<br>(3.00, 3.00) |
| How easy was it for you to use the CUE1 device in the recommended body position (e.g., sternum)?<sup>a</sup> | 3.00<br>(3.00, 3.00) |
| How easy was it for you to apply the adhesive patches provided with the CUE1 device to the recommended body position (e.g., sternum)?<sup>a</sup> | 3.00<br>(3.00, 3.00) |
| How likely are you to recommend the CUE1 device to others with Orthostatic Tremor who experience similar symptoms to yours?<sup>a</sup> | 3.00<br>(2.00, 3.00) |

Data are mean (SD) or median (IQR). N, number; %, percentage; SD= Standard deviation.
IQR= 1<sup>st</sup> and 3<sup>rd</sup> Interquartile. <sup>a</sup>Responses are shown on a 5-point Likert scale, where 0
indicates the lowest (e.g., not at all) rating and 4 indicates the highest (e.g., extremely) rating.

